# Bayesian analysis of tests with unknown specificity and sensitivity

**DOI:** 10.1101/2020.05.22.20108944

**Authors:** Andrew Gelman, Bob Carpenter

## Abstract

When testing for a rare disease, prevalence estimates can be highly sensitive to uncertainty in the specificity and sensitivity of the test. Bayesian inference is a natural way to propagate these uncertainties, with hierarchical modeling capturing variation in these parameters across experiments. Another concern is the people in the sample not being representative of the general population. Statistical adjustment cannot without strong assumptions correct for selection bias in an opt-in sample, but multilevel regression and poststratification can at least adjust for known differences between the sample and the population. We demonstrate hierarchical regression and poststratification models with code in Stan and discuss their application to a controversial recent study of SARS-CoV-2 antibodies in a sample of people from the Stanford University area. Wide posterior intervals make it impossible to evaluate the quantitative claims of that study regarding the number of unreported infections. For future studies, the methods described here should facilitate more accurate estimates of disease prevalence from imperfect tests performed on non-representative samples.

## 1. Background

Correction of diagnostic tests for false positives and false negatives is a well-known probability problem. When the base rate is low, estimates become critically sensitive to misclassifications (Hemenway, 1997). This issue hit the news recently (Lee, 2020), with a study of coronavirus antibodies in a population with a low incidence rate.

This is a problem where not fully accounting for uncertainty can make a big difference in scientific conclusions and potential policy recommendations. In early April, 2020, Bendavid et al. (2020a) recruited 3330 residents of Santa Clara County, California and tested them for SARS-CoV-2 antibodies. 50 people tested positive, yielding a raw estimate of 1.5%. After adjusting for differences between sample and population in sex, ethnicity, and zip code distributions, Bendavid et al. (2020a) reported an uncertainty range of 2.5% to 4.2%, implying that the number of infections in the county was between 50 and 85 times the count of cases reported at the time. Using an estimate of the number of coronavirus deaths in the county up to that time, they computed an implied infection fatality rate (IFR) of 0.12–0.2%, much lower than IFRs in the range of 0.5%–1% that had been estimated from areas with outbreaks of the disease.

The estimates from Bendavid et al. (2020a) were controversial, and it turned out that they did not correctly account for uncertainty in the specificity (true negative rate) of the test. There was also concern about the adjustment they performed for non-representativeness of their sample. Thus, the controversy arose from statistical adjustment and assessment of uncertainty. A revised preprint (Bendavid et al., 2020b) addressed some but not all of the problems with the analysis. It is possible that the authors of that study will prepare another analysis for eventual publication.

In the present article we set up a Bayesian framework to clarify these issues, specifying and fitting models using the probabilistic programming language Stan (Carpenter et al., 2017; Stan Development Team, 2020). There is a long literature on Bayesian measurement-error models (see Gustafson, 2003) and their application to diagnostic testing (Greenland, 2009); our contribution here is to set up the model, supply code, and consider multilevel regression and poststratification, influence of hyperpriors, and other challenges that arise in the problem of estimating population prevalence using test data from a sample of people.

## 2. Modeling a test with uncertain sensitivity and specificity

Testing for a rare disease is a standard textbook example of conditional probability, famous for the following counterintuitive result. Suppose a person tests positive for a disease, based on a test that has a 95% accuracy rate, and further suppose that this person is sampled at random from a population with a 1% prevalence rate. Then what is the probability that he or she actually has the disease? The usual intuition suggests that the conditional probability should be approximately 95%, but it is actually much lower, as can be seen from a simple calculation of base rates, as suggested by Gigerenzer et al. (2007). Imagine you test 1000 people. With a 1% prevalence rate, we can expect that 10 have the disease and 990 do not. Then, with a 95% accuracy rate (assuming this applies to both specificity and sensitivity of the test), we would expect 0.95 × 10 = 9.5 true positives and 0.05 × 990 = 49.5 false positives; thus, the proportion of positive tests that are true positives (i.e., the positive predictive value) is 9.5*/*(9.5 + 49.5) = 0.16, a number that is difficult to make sense of without visualizing the hypothetical populations of true positive and false positive tests.

A related problem is to estimate the prevalence of the disease given the rate of positive tests. If the population prevalence is *π* and the test has a specificity of *γ* and a sensitivity of *δ*, then the expected frequency of positive tests *p* is

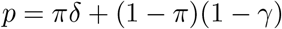

Given known *γ, δ* and *p*, we can solve for the prevalence,

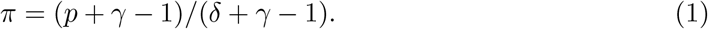

If the properties of the test are known, but *p* is estimated from a random sample, we can obtain a simple classical estimate by starting with a confidence interval for *p* and then propagating it through the formula. For example, Bendavid et al. (2020) report 50 positive tests out of 3330, which corresponds to an estimate 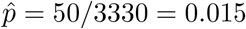 with standard error 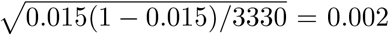. Supposing that their test had a specificity of *γ* = 0.995 and a sensitivity of *δ* = 0.80, this yields an estimate of (0.015+0.995*−*1)*/*(0.80+0.995*−*1) = 0.013 with standard error 0.002*/*(0.80 + 0.995 *−* 1) = 0.003.

Two immediate difficulties arise with the classical approach. First, if the observed rate 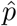 is less than 1 *− γ*, the false positive rate of the test, then the estimate from (1) becomes meaninglessly negative. Second, if there is uncertainty in the specificity and sensitivity parameters, it becomes challenging to propagate uncertainty through the nonlinear expression (1).

We can resolve both these problems with a simple Bayesian analysis (Gelman, 2020). First, suppose that priors for sensitivity and specificity have been externally supplied. The model is then

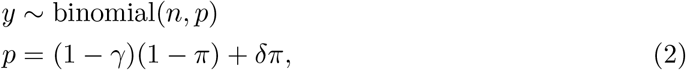

along with the specified prior distribution, *p*(*γ, δ*). In this model, the parameters *π, γ*, and *δ* must be constrained to be between 0 and 1, and *π* must be given a prior distribution too. A natural starting point would be *π* ∼ uniform(0, 1). In this case, previously existing knowledge of the population prevalence was weak enough that a reasonable prior on *π* should not have much impact on the posterior. The three parameters *π, γ*, and *δ*, are not jointly identified from only the number of positive test cases, hence the need for an informative prior on *γ* and *δ*. This can be seen as a generalization of the usual approach of assuming that these parameters are known exactly.

In the example of Bendavid (2020a), prior information on specificity and sensitivity was given in the form of previous trials, specifically *y*_*γ*_ negative results in *n*_*γ*_ tests of known negative subjects and *y*_*δ*_ positive results from *n*_*δ*_ tests of known positive subjects. This yields the model,

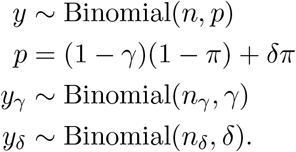

We use uniform(0, 1) priors on prevalence *π*, specificity *γ*, and sensitivity *δ*, with the understanding that they represent placeholders and could be augmented to include additional information. Stan code is in Appendix A.1.

We fit the model using the data reported in Bendavid et al. (2020a):

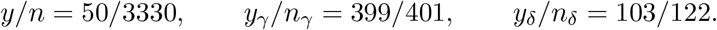

This results in high posterior uncertainty for the prevalence, *π*. Figure 1a shows the joint posterior simulations for *π* and *γ*: uncertainty in the population prevalence is in large part driven by uncertainty in the specificity. Figure 1b shows the posterior distribution for *π*, which reveals that the data and model are consistent with prevalences as low as 0% and as high as 2%.

**Figure 1:**
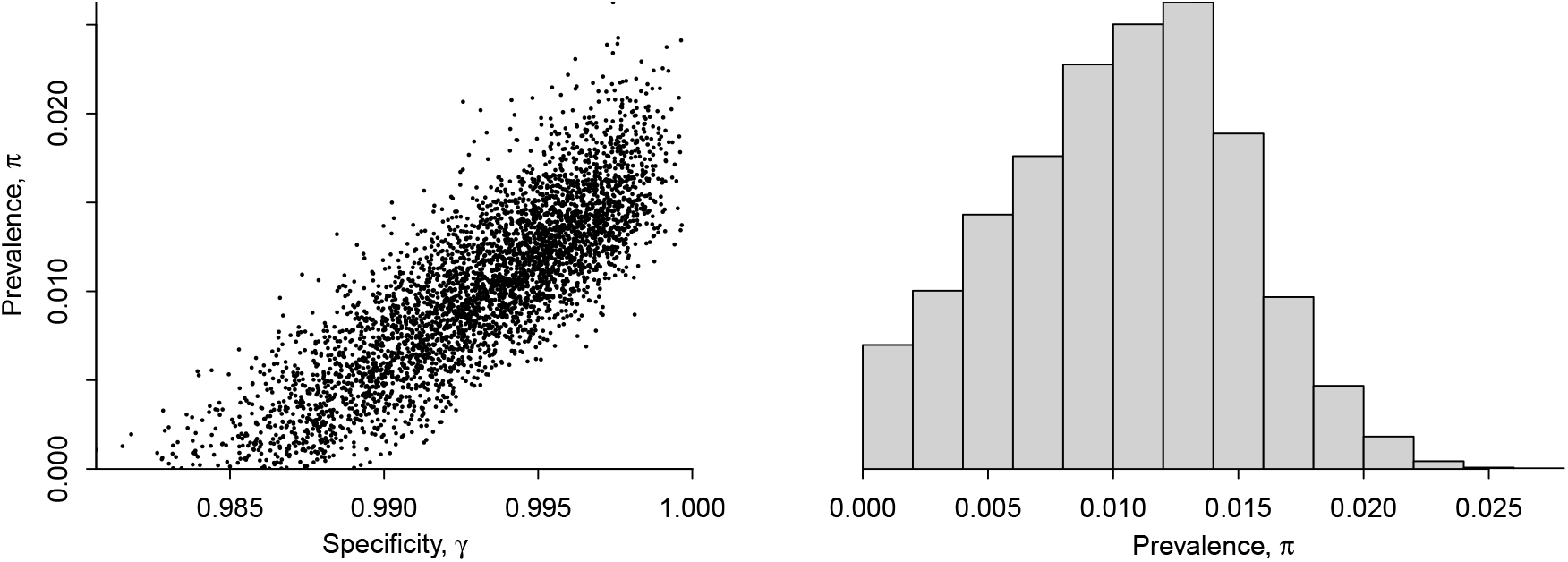
Summary of inference from model with unknown specificity, sensitivity, and prevalence, based on data from Bendavid et al. (2020a): (a) scatterplot of posterior simulations of prevalence, π, and specificity, γ; (b) histogram of posterior simulations of γ. This model assumes the testing sites are identical and thus pools all data.

The asymmetric posterior distribution with its hard bound at zero suggests that the usual central 95% interval will not be a good inferential summary. Instead we the use the shortest posterior interval^1^ for reasons discussed in Liu, Gelman, and Zheng (2015). The resulting 95% interval for *π* is (0, 1.8%), which is much different from the intervals reported by Bendavid et al. (2020a,b), with or without their correction for nonrepresentativeness of the sample. As a result, the substantive conclusion from that earlier report has been overturned. From the given data, the uncertainty in the specificity is large enough that the data do not supply strong evidence of a substantial prevalence.

## 3. Hierarchical model for varying testing conditions

The above analysis reveals that inference about specificity is key to precise estimation of low prevalence rates. In the second version of their report, Bendavid et al. (2020b) include data from 13 specificity studies and 3 sensitivity studies. Sensitivity and specificity can vary across experiments, so it is not appropriate to simply pool the data from these separate studies; indeed, these particular data are not consistent with constant error rates (Fithian, 2020). We allow the parameters to vary according to a hierarchical model where, for any study *j*, the specificity *γ*_*j*_ and sensitivity *δ*_*j*_ are drawn from normal distributions on the log odds (or logistic) scale,^2^

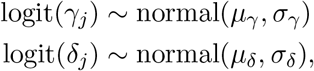

with the hyperparameters *µ* and *σ* can be estimated from the data. Stan code is given in Appendix A.2. In general it could make sense to allow correlation between *γ*_*j*_ and *δ*_*j*_ (Guo, Riebler, and Rue, 2017), but the way the data are currently available to us, specificity and sensitivity are estimated from separate studies and so there is no information about such a correlation. When coding the model, we use the convention that *j* = 1 corresponds to the study of interest, with other *j >* 1 representing studies of specificity or sensitivity given known samples. The parameters *γ*_1_ and *δ*_1_ represent the specificity and sensitivity for the site performing the prevalence study (the 50/3330 positive tests of patients with unknown status).

One could also consider alternatives to the logistic transform, which allows the unbounded normal distribution to map to the unit interval but might not be appropriate for tests where the specificity can actually reach the value of 1.

We fit the above hierarchical model to the data from Bendavid et al. (2020b), assigning a uniform prior to *π* and weak normal^+^(0, 1) priors to *σ*_*γ*_, *σ*_*δ*_ (using the notation normal^+^ for the truncated normal distribution constrained to be positive). We often use half-normal or half-*t* priors for variance parameters when we want to constrain them at the high end but allow them to be arbitrarily close to zero if the data support such inferences (Gelman, 2006). Setting the scale of these half-normals to 1 makes the prior weak for this particular application, in the following sense. A shift of 1 on the logit scale represents a pretty big change in sensitivity or specificity. For example, logit(0.8) = 1.4, so if 0.8 is a typical value of sensitivity, and if *σ*_*δ*_ = 1, then we would expect sensitivities to vary by roughly *±*1 standard deviation, or 0.4 to 2.4 on the logit scale, which corresponds to a probability range from 0.60 to 0.92. The normal^+^(0, 1) hyperpriors weakly pull the specificities and sensitivities from different studies toward each other, while allowing for a large variation if required by the data.

The resulting posterior inference is shown in Figure 2a. The 95% posterior interval for the prevalence is now (0.000, 0.160). Where does that upper bound come from: how could an underlying prevalence of 16% be plausible, given that only 1.5% of the people in the sample tested positive? The answer can be seen from the large uncertainty in the sensitivity parameter, which in turn comes from the possibility that *σ*_*δ*_ is very large. The trouble is that the sensitivity information in these data comes from only three experiments, which is not enough to get a good estimate of the underlying distribution. This problem is discussed by Guo, Riebler, and Rue (2017).

**Figure 2:**
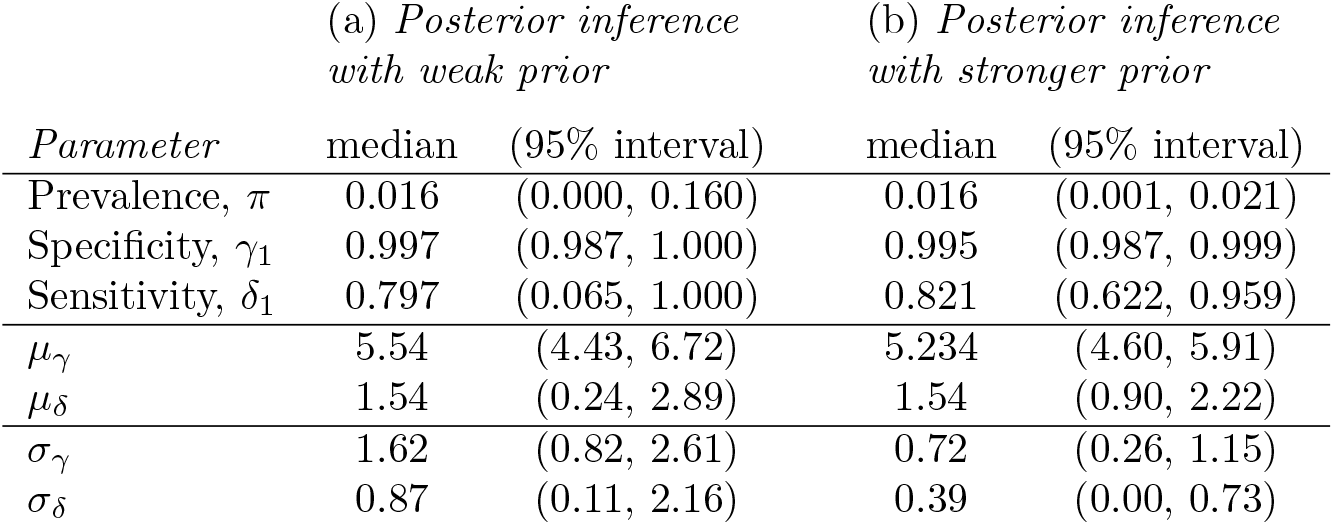
Summary of inferences (posterior median and shortest 95% posterior interval) for the prevalence, specificity, and sensitivity of the Bendavid et al. (2020b) study, along with inferences for the hyperparameters characterizing the distribution of specificity and sensitivity on the logistic scale. (a) For the model with weak priors for σ_γ_ and σ_δ_, the posterior inference for the prevalence, π, is highly uncertain. This is driven by uncertainty in the sensitivity, which in turn is driven by uncertainty in the hyperparameters for the sensitivity distribution. (b) Stronger priors on σ_γ_ and σ_δ_ have the effect of regularizing the specificity and sensitivity parameters, leading to narrower intervals for π, the parameter of interest in this study. The hyperparameters µ and σ are on the logistic scale and thus are difficult to interpret without transformation.

The only way to make progress here is to constrain the sensitivity parameters in some way. One possible strong assumption is to assume that *σ*_*δ*_ is some small value. This could make sense in the current context, as we can consider it as a relaxation of the assumption of Bendavid et al. (2020b) that *σ*_*δ*_ = 0. We also have reason to believe that specificity will not vary much between experiments, so we will apply a soft constraint to the variation in specificities as well.

Instead of specifying *σ*_*δ*_, we give it an informative prior distribution. In particular, we replace the weakly informative normal^+^(0, 1) priors on *σ*_*γ*_, *σ*_*δ*_ with something stronger, *σ*_*γ*_, *σ*_*δ*_ *∼* normal^+^(0, 0.3). To get a sense of what this means, start with the point estimate from Figure 2a of *µ*_*δ*_, which is 1.58. If *σ*_*δ*_ were 0.3, then there would be a roughly 2/3 chance that the sensitivity of in a new experiment is in the range logit^*−*1^(1.58 *±* 0.3), which is (0.78, 0.87). This seems reasonable.

Figure 2b shows the results. Our 95% interval for *π* is now (0.001, 0.021); that is, the infection rate is estimated to be somewhere between 0.1% and 2.1%.

## 4. Prior sensitivity analysis

To assess the sensitivity of the above prevalence estimate to the priors placed on *σ*_*γ*_ and *σ*_*δ*_, we consider the family of prior distributions,

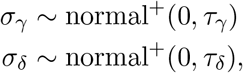

where *τ*_*δ*_ and *τ*_*γ*_ are user-specified hyperparameters. Setting *τ*_*δ*_ and *τ*_*γ*_ to zero would force *σ*_*δ*_ and *σ*_*γ*_ to be zero and would enforce complete pooling, corresponding to Bendavid et al.’s (2020b) assumption that each test site has identical specificity and sensitivity. As the hyperparameters are increased, the scales of variation of *σ*_*γ*_ and *σ*_*δ*_ are allowed to vary more, and setting *τ*_*γ*_ and *τ*_*δ*_ to infinity would typically be considered noninformative in the sense of providing the least amount of constraint on the sensitivities and specificities. In practice, we often use normal^+^(0, 1) priors for hierarchical scale parameters, on the default assumption that the underlying parameters (in this case, the specificities) will probably vary by less than 1 on the logit scale.

For this problem, however, a weak prior does not work: as shown in the left panel of Figure 2, the resulting inferences for the sensitivities are hopelessly wide. We do not believe these tests have specificities below 50%, yet such a possibility is included in the posterior distribution, and this in turn propagates to inappropriately wide intervals for the prevalence, *π*. As explained in the previous section, that is why we assigned a stronger prior, using hyperprior parameters *τ*_*γ*_ = *τ*_*δ*_ = 0.2.

Figure 3 shows how these hyperprior parameters *τ*_*γ*_ and *τ*_*δ*_ affect inferences for the prevalence, *π*. The posterior median of *π* is not sensitive to the scales *τ*_*γ*_ and *τ*_*δ*_ of the hyperpriors, but the uncertainty in that estimate, as indicated by the central posterior 90% intervals, is influenced by these settings. In particular, in the graphs on the right, when the sensitivity hyperprior parameter *τ*_*δ*_ is given a high value, the upper end of the interval is barely constrained. The lower end of the interval is fairly stable, as long as the specificity hyperprior parameter *τ*_*γ*_ is not given an artificially low value. Here we are using central rather than shortest posterior intervals because we are displaying inference on the log scale and so there is no boundary.

**Figure 3:**
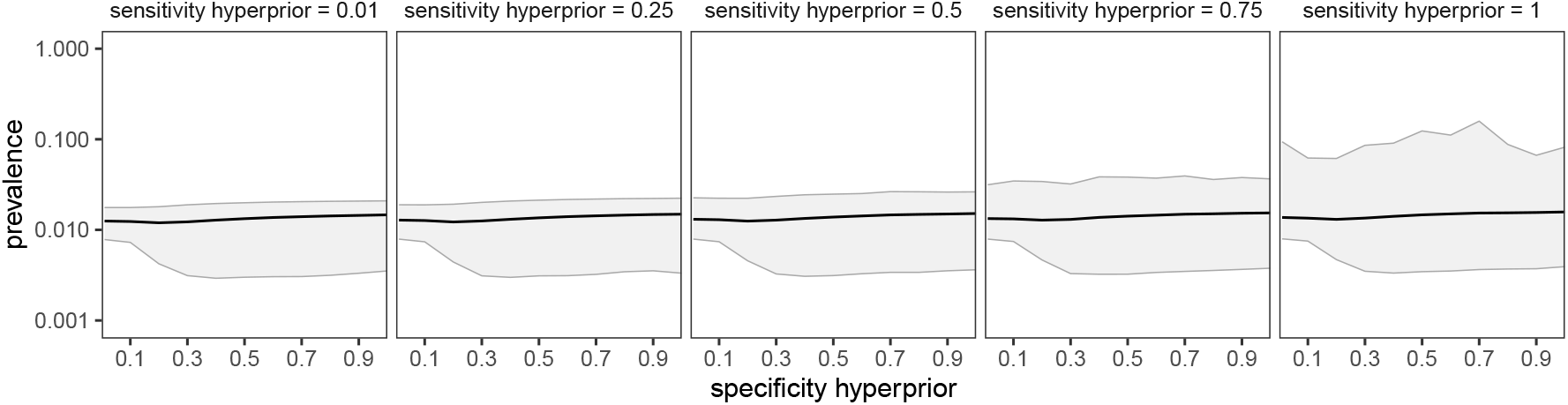
Each panel shows a plot of the posterior median and central 90% posterior interval of the prevalence, π, as a function of τ_γ_ and τ_δ_, the prior scales for the specificity and sensitivity hyperparameters, σ_γ_ and σ_δ_. The posterior median of prevalence is not sensitive to τ_γ_ and τ_δ_, but the endpoints of the 90% interval show some sensitivity. It is possible to use a weak hyperprior on the scale of the specificity distribution, σ_γ_: this makes sense given that there are 13 prior specificity studies in the data. For the scale of the sensitivity distribution, σ_δ_, it is necessary to use a prior scale of 0.5 or less to effectively rule out the possibility of extremely high prevalence corresponding to an unrealistic sensitivity parameter γ. The noise in the rightmost graph represents Monte Carlo error that is a consequence of the weakly specified model.

When *τ*_*γ*_ and *τ*_*δ*_ are too low, the variation in specificity and sensitivity are constrained to be nearly zero, all values are pooled, and uncertainty is artificially deflated. As the hyperprior parameters are increased, the uncertainty in prevalence increases. This gets out of hand when the hyperprior for sensitivity is increased, because there are only three data points to inform the distribution it controls. This is an example of the general principle that wide hyperpriors on hierarchical scale parameters can pull most of the probability mass into areas of wide variation and dominate the data, leading to inflated uncertainty. Around the middle of these ranges, the posterior intervals are not as sensitive to variation in the hyperpriors. We would consider values *τ*_*γ*_ = *τ*_*δ*_ = 0.5 to be weakly informative for this example, in that they are roughly consistent with inter-site variation in specificity in the range 73% to 99.3% and of specificity in the range 88% to 99.75%.

In addition we need priors on *µ*_*γ*_ and *µ*_*δ*_. In this particular example, once we have constrained the variation in the specificities and sensitivities, enough data are available to estimate these population means with uniform priors on these parameters, but in general it is best to use prior information to at least roughly constrain them. For this example, we assign independent normal(4, 2) priors, a distribution that puts 2/3 of its mass in the range 4 *±* 2, which, after undoing the logistic transformation, corresponds to (0.881, 0.997) on the probability scale, which seem like a suitably broad range for the mean of the population distribution of specificity and sensitivity of these tests.

The complexity of this sensitivity analysis might seem intimidating: if Bayesian inference is this difficult and this dependent on priors, maybe it is not a good idea?

We would argue that the problem is not as difficult as it might look. The steps taken in Sections 2 and 3 show the basic workflow: We start with a simple model, then add hierarchical structure. For the hierarchical model we started with weak priors on the hyperparameters and examined the inferences, which made us realize that we had prior information (that specificities and sensitivities of the tests should not be so variable), which we then incorporated into the next iteration of the model. Performing the sensitivity analysis was fine—it helped us understand the inferences better—but it was not necessary for us to get reasonable inferences.

Conversely, non-Bayesian analyses would not be immune from this sensitivity to model choices, as is illustrated by the mistakes made by Bendavid et al. (2020b) to treat specificity and sensitivity as not varying at all, to set *σ*_*γ*_ = *σ*_*δ*_ = 0 in our notation. An alternative could be to use the calibration studies to get point estimates of *σ*_*γ*_ and *σ*_*δ*_, but then there would still be the problem of accounting for uncertainty in these estimates, which would return the researchers to the need for some sort of external constraint or bound on the distribution of the sensitivity parameters *δ*_*j*_, given that only three calibration studies are available here to estimate these. This in turn suggests the need for more data or modeling of the factors that influence the test’s specificity and sensitivity. In short, the analysis shown in Figure 3 formalizes a dependence on prior information that would arise, explicitly or implicitly, in any reasonable analysis of these data.

## 5. Extensions of the model

### 5.1. Multilevel regression and poststratification (MRP) to adjust for differences between sample and population

Bendavid et al. (2020a,b) compared demographics on 3330 people they tested, and they found differences in the distributions of sex, age, ethnicity, and zip code of residence compared to the general population of Santa Clara County. It would be impossible to poststratify the raw data on 2 sexes, 4 ethnicity categories, 4 age categories, and 58 zip codes, as the resulting 1856 cells would greatly outnumber the positive tests in the data. They obtained population estimates by adjusting for sex × ethnicity × zip code, but their analysis is questionable, first because they did not adjust for age, and second because of noisy weights arising from the variables they did adjust for. To obtain stable estimates while adjusting for all these variables, we would recommend applying a multilevel model to the exposure probability, thus replacing the constant *π* in the above models with something like the following logistic regression.^3^

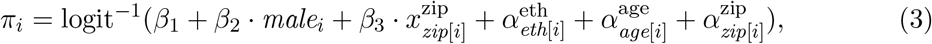

where *male* is a variable that takes on the value 0 for women and 1 for men; *x*^zip^ is a relevant predictor at the zip code level; *eth*[*i*], *age*[*i*], and *zip*[*i*] are index variables for survey respondent *i*; the *β* parameters are logistic regression coefficients; and the *α* parameters are vectors of varying intercepts. These varying intercepts have hierarchical priors

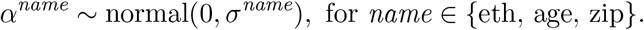

In the regression model (3), it is important to include the predictor *x*^zip^, which in this example might be percent Latino or average income in the zip code. Otherwise, with so many zip codes, the multilevel model will just partially pool most of the zip code adjustments to zero, and not much will be gained from the geographic poststratification. The importance of geographic predictors is well known in the MRP literature; see, for example, (Caughey and Warshaw 2019).

In addition, priors are needed for *σ*^eth^, *σ*^age^, *σ*^zip^, and *β*, along with the hierarchical specificity and sensitivity parameters from the earlier model. For these hyperparameters, we assign normal^+^(0, 0.5) priors for *σ*^eth^, *σ*^age^, and *σ*^zip^. These priors allow the prevalence to vary moderately by these poststratification factors.

We use a unit logistic prior for the centered intercept 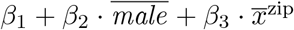 (corresponding to a uniform(0, 1) prior distribution for the probability that an average person in the sample has the antibody), a normal(0, 0.5) prior for *β*_2_, and a normal(0, 0.5*/s*_zip_) for *β*_3_, where *s*_zip_ is the standard deviation of *x*_zip_ in the data.

The point of the scaling of *β*_2_ and *β*_3_ is to give some prior regularization on the contribution of each predictor in the data. Regarding the prior on the intercept: Stan allows direct assignment of distributions to transformed parameters; in this particular case, the transform is affine and thus does not require a Jacobian adjustment. By assigning prior distributions to the centered intercept and two other regression coefficients, we have implicitly assigned a prior distribution to the three parameters, (*β*_1_, *β*_2_, *β*_3_).

We code the model in Stan; see Appendix A.3. Unfortunately the raw data from Ben-david et al. are not currently available, so we fit the model to simulated data to check the stability of the computation.

The above model is a start; it could be improved by including interactions, following the general ideas of Ghitza and Gelman (2013). In any case, once this model has been fit, it can be used to make inferences for disease prevalence for all cells in the population. As discussed by Johnson (2020), these cell estimates can then be summed, weighting by known population totals (in this case, the number of people in each sex × ethnicity × age × zip code category in the population) to get inferences for the prevalence in the county,

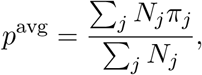

where *N*_*j*_ is the number of people in cell *j* in the general population, and *π*_*j*_ is the prevalence in cell *j* as computed from the logistic model. We perform this summation in the generated quantities block of the Stan model in Appendix A.3.

### 5.2 Variation across location and over time

The aforementioned Santa Clara County study is just one of many recent SARS-CoV-2 antibody surveys. Other early studies were conducted in Boston, New York, Los Angeles, and Miami, and in various places outside the United States, and we can expect many more in the future. If the raw data from these studies were combined, it should be possible to estimate the underlying prevalences from all these studies using a hierarchical model, allowing specificity, sensitivity, and prevalence to vary by location, and adjusting for non-sampling error where possible. Such an analysis is performed by Levesque and Maybury (2020) using detailed information on the different tests used in different studies.

We will also be seeing more studies of changing infection rates over time. Stringhini et al. (2020) perform such an analysis of weekly surveys in Geneva, Switzerland, accounting for specificity and sensitivity and poststratifying by sex and age.

### 5.3 Including additional diagnostic data

We have so far assumed that test results are binary, but additional information can be gained from continuous measurements that make use of partial information when data are near detection limits (Gelman, Chew, and Shnaidman, 2004; Bouman, Bonhoeffer, and Regoes, 2020). Further progress can be made by performing different sorts of tests on study participants or retesting observed positive results.

Another promising direction is to include additional information on people in the study, for example from self-reported symptoms. Some such data are reported in Bendavid et al. (2020b), although not at the individual level. With individual-level symptom and test data, a model with multiple outcomes could yield substantial gains in efficiency compared to the existing analysis using only a single positive/negative test result on each participant.

A third direction would be to acquire test results from sites testing both known positive and known negative cases. With such tests, bivariate priors for sensitivity and specificity could be formulated as suggested by Guo, Riebler, and Rue (2017). Simple multivariate normal priors are possible, but the situation is complicated because, in general, sensitivity is negatively correlated with specificity in diagnostic tests, but above or below average testing quality at the sites will provide positive correlation. Thus it may be better to formulate priors in terms of bias (trading sensitivity for specificity) and accuracy instead of directly in terms of sensitivity and specificity.

## 6. Non-Bayesian approaches

As with any statistical analysis, alternative approaches are possible that would use the same information and give similar results.

In Section 2, it was necessary to account for uncertainty in all three parameters, while respecting the constraint that all three probabilities had to be between 0 and 1. We assume that both these aspects of the model could be incorporated into a non-Bayesian approach by working out the region in the space of (*π, γ, δ*) that is consistent with the data and then constructing a family of tests which could be inverted to create confidence regions.

This could be expanded into a multilevel model as in Section 3 by considering the specificities and sensitivities of the different experiments as missing data and averaging over their distribution, but still applying non-Bayesian inference to the resulting hyperparameters. The wide uncertainty intervals from the analysis in Section 3 suggest that some constraints or regularization or additional information on the hyperparameters would be necessary to get stable inferences here, no matter what statistical approach is used.

Fithian (2020) performs a non-Bayesian analysis of the data from Bendavid et al. (2020b), coming to the same basic conclusion that we do, demonstrating that the calibration data are incompatible with a model of constant specificity and that, once the specificity is allowed to vary, the observed rate of positive tests in the Santa Clara study does not allow rejection of the null hypothesis of zero infection rate. Had it been possible to reject zero, this would not be the end of the story: at that point one could invert a family of tests to obtain a confidence region, as noted above.

Finally, some rough equivalent to the poststratification adjustment in Section 5.1 could be performed using a non-Bayesian weighting approach, using some smoothing to avoid the noisiness of raw poststratification weights. Similarly, non-Bayesian methods could be used to fit regressions allowing prevalence to vary over location and time.

## 7. Discussion

### 7.1. Limitations of the statistical analysis

Epidemiology in general, and disease testing in particular, feature latent parameters with high levels of uncertainty, difficulty in measurement, and uncertainty about the measurement process as well. This is the sort of setting where it makes sense to combine information from multiple studies, using Bayesian inference and hierarchical models, and where inferences can be sensitive to assumptions.

The biggest assumptions in this analysis are, first, that the historical specificity and sensitivity data are relevant to the current experiment; and, second, that the people in the study are a representative sample of the general population. We addressed the first concern with a hierarchical model of varying sensitivities and specificities, and we addressed the second concern with multilevel regression and poststratification on demographics and geography. But this modeling can take us only so far. If there is hope or concern that the current experiment has unusual measurement properties, or that the sample is unrepresentative in ways not accounted for in the regression, then more information or assumptions need to be included in the model, as described by Campbell et al. (2020).

The other issue is that there are choices of models, and tuning parameters within each model. Sensitivity to the model is apparent in Bayesian inference, but it would arise with any other statistical method as well. For example, Bendavid et al. (2020a) used an (incorrectly applied) delta method to propagate uncertainty, but this is problematic when sample size is low and probabilities are near 0 or 1. Bendavid et al. (2020b) completely pooled their specificity and sensitivity experiments, which is equivalent to setting *σ*_*γ*_ and *σ*_*δ*_ to zero. And their weighting adjustment has many arbitrary choices. We note these not to single out these particular authors but rather to emphasize that, at least for this problem, all statistical inferences involve user-defined settings.

For the models in the present article, the most important user choices are: (a) what data to include in the analysis, (b) prior distributions for the hyperparameters, and (c) the structure and interactions to include in the MRP model. For these reasons, it would be difficult to set up the model as a plug-and-play system where users can just enter their data, push a button, and get inferences. Some active participation in the modeling process is required, which makes sense given the sparseness of the data. When studying populations with higher prevalences and with data that are closer to random samples, more automatic approaches might be possible.

### 7.2 Santa Clara study

Section 3 shows our inferences given the summary data in Bendavid et al. (2020b). The inference depends strongly on the priors on the distributions of sensitivity and specificity, but that is unavoidable: the only way to avoid this influence of the prior would be to sweep it under the rug, for example by just assuming a zero variation in the test parameters.

What about the claims regarding the rate of coronavirus exposure and implications for the infection fatality rate? It is hard to say from this one study: the numbers in the data are consistent with zero infection rate and a wide variation in specificity and sensitivity across tests, and the numbers are also consistent with the claims made in Bendavid et al. (2020a,b). That does not mean anyone thinks the true infection rate is zero. It just means that more data, assumptions, and subject-matter knowledge are required. That is to be expected—people usually make lots of assumptions in analyzing this sort of laboratory assay. It is common practice to use the manufacturer’s numbers on specificity, sensitivity, detection limit, and so forth, and not worry about that level of variation. Only when estimating a very low underlying rate do the statistical challenges become so severe. This is an example of the general phenomenon in statistics that the severity of identification problems can depend on the data.

For now, we do not think the data support the claim that the number of infections in Santa Clara County was between 50 and 85 times the count of cases reported at the time, or the implied interval for the IFR of 0.12–0.2%. These numbers are consistent with the data, but the data are also consistent with a near-zero infection rate in the county. The data of Bendavid et al. (2020a,b) do not provide strong evidence about the number of people infected or the infection fatality ratio; the number of positive tests in the data is just too small, given uncertainty in the specificity of the test.

The analyses in this article suggest that future studies should be conducted with full awareness of the challenges of measuring specificity and sensitivity, that relevant variables be collected on study participants to facilitate inference for the general population, and that (de-identified) data be made accessible to external researchers.

## Data Availability

All data and code for this paper are freely available

https://bob-carpenter.github.io/diagnostic-testing/

## A. Stan programs

### A.1. Model with binomial data on specificity and sensitivity

~~~
data {
 int<lower = 0> y_sample;
 int<lower = 0> n_sample;
 int<lower = 0> y_spec;
 int<lower = 0> n_spec;
 int<lower = 0> y_sens;
 int<lower = 0> n_sens;
}
parameters {
 real<lower=0, upper = 1> p;
 real<lower=0, upper = 1> spec;
 real<lower=0, upper = 1> sens;
}
model {
 real p_sample = p * sens ∗ (1 - p) ∗ (1 - spec);
 y_sample ∼ binomial(n_sample, p_sample);
 y_spec ∼ binomial(n_spec, spec);
 y_sens ∼ binomial(n_sens, sens);
}
~~~

### A.2. Hierarchical model for specificities and sensitivities

~~~
data {
 int<lower = 0> y_sample;
 int<lower = 0> n_sample;
 int<lower = 0> J_spec;
 int<lower = 0> y_spec[J_spec];
 int<lower = 0> n_spec[J_spec];
 int<lower = 0> J_sens;
 int<lower = 0> y_sens[J_sens];
 int<lower = 0> n_sens[J_sens];
 real<lower = 0> logit_spec_prior_scale;
 real<lower = 0> logit_sens_prior_scale;
}
parameters {
 real<lower = 0, upper = 1> p;
 real mu_logit_spec;
 real mu_logit_sens;
 real<lower = 0> sigma_logit_spec;
 real<lower = 0> sigma_logit_sens;
 vector<offset = mu_logit_spec, multiplier = sigma_logit_spec>[J_spec] logit_spec;
 vector<offset = mu_logit_sens, multiplier = sigma_logit_sens>[J_sens] logit_sens;
}
transformed parameters {
 vector[J_spec] spec = inv_logit(logit_spec);
 vector[J_sens] sens = inv_logit(logit_sens);
}
model {
 real p_sample = p * sens[1] + (1 - p) * (1 - spec[1]);
 y_sample ∼ binomial(n_sample, p_sample);
 y_spec ∼ binomial(n_spec, spec);
 y_sens ∼ binomial(n_sens, sens);
 logit_spec ∼ normal(mu_logit_spec, sigma_logit_spec);
 logit_sens ∼ normal(mu_logit_sens, sigma_logit_sens);
 sigma_logit_spec ∼ normal(0, logit_spec_prior_scale);
 sigma_logit_sens ∼ normal(0, logit_sens_prior_scale);
 mu_logit_spec ∼ normal(4, 2); // weak prior on mean of distribution of spec
 mu_logit_sens ∼ normal(4, 2); // weak prior on mean of distribution of sens
}
~~~

### A.3 Multilevel regression and poststratification

~~~
data {
 int<lower = 0> N; // number of tests in the sample (3330 for Santa Clara)
 int<lower = 0, upper = 1> y[N]; // 1 if positive, 0 if negative
 vector<lower = 0, upper = 1>[N] male; // 0 if female, 1 if male
 int<lower = 1, upper = 4> eth[N]; // 1=white, 2=asian, 3=hispanic, 4=other
 int<lower = 1, upper = 4> age[N]; // 1=0-4, 2=5-18, 3=19-64, 4=65+
 int<lower = 0> N_zip; // number of zip codes (58 in this case)
 int<lower = 1, upper = N_zip> zip[N]; // zip codes 1 through 58
 vector[N_zip] x_zip; // predictors at the zip code level
 int<lower = 0> J_spec;
 int<lower = 0> y_spec [J_spec];
 int<lower = 0> n_spec [J_spec];
 int<lower = 0> J_sens;
 int<lower = 0> y_sens [J_sens];
 int<lower = 0> n_sens [J_sens];
 int<lower = 0> J; // number of population cells, J = 2*4*4*58
 vector<lower = 0>[J] N_pop; // population sizes for poststratification
 real<lower = 0> coef_prior_scale;
 real<lower = 0> logit_spec_prior_scale;
 real<lower = 0> logit_sens_prior_scale;
}
parameters {
 real mu_logit_spec;
 real mu_logit_sens;
 real<lower = 0> sigma_logit_spec;
 real<lower = 0> sigma_logit_sens;
 vector<offset = mu_logit_spec, multiplier = sigma_logit_spec>[J_spec] logit_spec;
 vector<offset = mu_logit_sens, multiplier = sigma_logit_sens>[J_sens] logit_sens;
 vector[3] b; // intercept, coef for male, and coef for x_zip
 real<lower = 0> sigma_eth;
 real<lower = 0> sigma_age;
 real<lower = 0> sigma_zip;
 vector<multiplier = sigma_eth>[4] a_eth; // varying intercepts for ethnicity
 vector<multiplier = sigma_age>[4] a_age; // varying intercepts for age category
 vector<multiplier = sigma_zip>[N_zip] a_zip; // varying intercepts for zip code
}
transformed parameters {
 vector[J_spec] spec = inv_logit(logit_spec);
 vector[J_sens] sens = inv_logit(logit_sens);
}
model {
 vector[N] p = inv_logit(b[1]
            + b[2] * male
            + b[3] * x_zip[zip]
            + a_eth[eth]
            + a_age[age]
            + a_zip[zip]);
 vector[N] p_sample = p * sens[1] + (1 - p) * (1 - spec[1]);
 y ∼ bernoulli(p_sample);
 y_spec ∼ binomial(n_spec, spec);
 y_sens ∼ binomial(n_sens, sens);
 logit_spec ∼ normal(mu_logit_spec, sigma_logit_spec);
 logit_sens ∼ normal(mu_logit_sens, sigma_logit_sens);
 sigma_logit_spec ∼ normal(0, logit_spec_prior_scale);
 sigma_logit_sens ∼ normal(0, logit_sens_prior_scale);
 mu_logit_spec ∼ normal(4, 2); // weak prior on mean of distribution of spec
 mu_logit_sens ∼ normal(4, 2); // weak prior on mean of distribution of sens a_eth ∼ normal(0, sigma_eth);
 a_age ∼ normal(0, sigma_age);
 a_zip ∼ normal(0, sigma_zip);
 // prior on centered intercept
 b[1] + b[2] * mean(male) + b[3] * mean(x_zip[zip]) ∼ logistic(0, 1);
 b[2] ∼ normal(0, coef_prior_scale);
 b[3] ∼ normal(0, coef_prior_scale / sd(x_zip[zip])); // prior on scaled coef
 sigma_eth ∼ normal(0, coef_prior_scale);
 sigma_age ∼ normal(0, coef_prior_scale);
 sigma_zip ∼ normal(0, coef_prior_scale);
}
generated quantities {
 real p_avg;
 vector[J] p_pop; // population prevalence in the J poststratification cells
 int count;
 count = 1;
 for (i_zip in 1:N_zip) {
  for (i_age in 1:4) {
   for (i_eth in 1:4) {
    for (i_male in 0:1) {
     p_pop[count] = inv_logit(b[1]
                + b[2] * i_male
                + b[3] * x_zip[i_zip]
                + a_eth[i_eth]
                + a_age[i_age]
                + a_zip[i_zip]);
     count += 1;
    }
   }
  }
 }
 p_avg = sum(N_pop .* p_pop) / sum(N_pop);
}
~~~

## Footnotes

1 The shortest posterior interval is equivalent to the highest posterior density interval for unimodal posteriors as we have here.

2 The log odds function is defined by logit(*p*) = log(*p/*(1 *− p*)).

3 *x*[*i*] and *x*_*i*_ are used interchangeably to improve readability.

